# A multi-centric, cross-sectional study on COVID-19 in Bangladesh: Clinical epidemiology and short-term outcomes in recovered individuals

**DOI:** 10.1101/2020.09.09.20191114

**Authors:** Adnan Mannan, H. M. Hamidullah Mehedi, Naim Uddin Hasan A Chy, Md. Omar Qayum, Farhana Akter, Abdur Rob, Prasun Biswas, Sanjida Hossain, Mustak Ibn Ayub

## Abstract

**Background:** COVID-19 turned into a global pandemic rapidly. This study was aimed to investigate SARS-CoV-2 associated epidemiology and clinical outcomes in Bangladesh in order to understand the future course of COVID-19 pandemic and develop prevention approaches.

**Design and Methods:** A cross-sectional retrospective study was conducted for a sample of 1,021 RT-PCR confirmed COVID-19 cases admitted in six different hospitals in Bangladesh and who recovered four weeks prior to the interview date.

**Results:** Of the total sample, 111 (10.9%) cases were asymptomatic while the number of symptomatic cases were 910 (89.1%). Higher prevalence of COVID-19 persisted in the male population (75%), cohorts having B (+) ve blood group (36.3%) and for the 31-40 age group. More than 85% of the sample reported a BCG vaccination mark. Common symptoms observed in our study samples were fever (72.4%), cough (55.9%), loss of taste (40.7%) and body ache (40%); whereas among the biochemical parameters, Neutrophil (46.4%), D-dimer (46.1%), and Ferritin (37.9%) levels were found elevated. Post-COVID complications including pain (31.8%), loss of concentration (24.4%) and anxiety or depression (23.1%) were also found significantly prevalent in the symptomatic cases with commodities.

**Conclusion:** Our study has shown that adult males aged in between 31-40 in Bangladesh are more vulnerable to being infected with COVID-19. The study also indicates a rising trend of the asymptomatic cases as the pandemic progresses deeper in time, and hence, deployment of interventions to curb further spread of community infection is necessary to avoid the grave outcomes of COVID-19 in Bangladesh.

## Introduction

The Coronavirus Disease 2019 or COVID-19 is a contagious disease of the respiratory system.^1^ In terms of genetic characteristics, SARS-CoV-2 is significantly diverse from SARS-CoV and MERS-CoV.^2, 3^ The virus’s rapid progression around the globe has unfolded a variety of clinical manifestations in different geographical locations. Severe prognosis of COVID-19 has been reported to be associated with comorbidities including diabetes, hypertension, cardiovascular disease, chronic obstructive pulmonary disease (COPD), malignancy, and chronic liver disease.^4^

With a population of more than 161 million people, Bangladesh stands eighth among the most populated countries in the world.^5^ In Bangladesh, as of 9 October, 2020, infections from SARS-CoV-2 has spiked to nearly 375,870 individuals while the death count figured to 5477 people (https://iedcr.gov.bd/). In regards to the clinical outcomes of COVID-19 and its association with various pathophysiological factors, no comprehensive study has been conducted in Bangladesh. As some of the studies were not conducted for a large sample size,^6-8^ understanding a bigger picture of the aforementioned relationship of COVID-19 symptoms with comorbidities and biochemical parameters from previous studies was not possible. Especially, the long-term complications among patients recovered from COVID-19 have not been conducted with necessary rigor and comprehension. Investigating the epidemiological characteristics of individuals diagnosed with COVID-19 in Bangladesh will help in providing a proper insight of the clinical characterization and patterns in progression of the disease. Therefore, it is vital to examine these aspects and factors related to the outcomes of COVID-19 for enacting appropriate means of prevention and treatment.

Since to this date, this study is a pioneering approach for the epidemiological characterization of COVID-19 individuals in Bangladesh, data related to the clinical aspects of COVID-19 is scarce, we managed to create a large sample size for our study by collecting data for a consecutive three months during the pandemic. This study aimed to investigate the patterns and array of symptoms in confirmed COVID-19 individuals as well as assess its relationship with the presence of comorbidities and results from biochemical assays with various other preliminary long-term clinical expositions post recovery.

## Methods

### Study design

A cross-sectional retrospective study was conducted among the cases confirmed to be COVID-19 positive following assessment by RT-PCR using any one of the following samples: throat swabs / nasal swabs /blood. These COVID-19 diagnosed patients were admitted in six different hospitals in Bangladesh. Scrutinization of post COVID-19 clinical characteristics was done upon each subject’s discharge from the hospital and after confirming that these subjects were indeed COVID-19 free by conducting two consecutive RT-PCR assays 24 hours apart. Recovered patients (according to RT-PCR negative result) at least four weeks before the interview were considered for this study. We also categorized all positive patients into two categories (symptomatic and asymptomatic) according to the presence of any one of the established symptoms referred by WHO/CDC.

### Study sites and sample size

The study was conducted in six hospitals having specialized units (following government instructions) for isolating and treating COVID-19 individuals from two different divisions of Bangladesh namely Dhaka (Dhaka Mohanogor General Hospital and Narayanganj Sadar Hospital) and Chattogram (Chittagong General Hospital, Chittagong Medical College Hospital, Chattogram Field Hospital and Chandpur Sadar Hospital). The study period started from 1 April 2020 and ended on 30 June 2020.

### Data collection and case enrollment

The physicians collected data relevant to the study retrospectively by interviewing 1021 COVID-19 patients over the telephone and all data were recorded into a preset Google Form. After double checking each information, the Google Forms were submitted and stored in our database. Medical prescriptions, hospital records were also accessed and matched with patients’ data obtained through the interview. Questionnaires included patient’s socio-demographic information, clinical manifestations, biochemical parameters, behavioral practice, comorbidities, medications, vitals, laboratory tests, electrocardiogram results, inpatient medications, treatments and outcomes (including length of stay, discharge, re-admission, and mortality).

### Exclusion criteria

In this study, deceased patients and those who were not interested to participate or did not give consent to data collection and usage for research purposes were not included.

### Ethical consideration

The protocol was approved by IRB of 250 bedded General Hospital, Chattogram, Bangladesh. The ethical approval number is 00981.

### Statistical analysis

Descriptive statistical analyses were performed expressing categorical variables with numbers and proportions. They were compared using chi-square test and Fisher’s exact test. P values of less than or equal to 0.05 (two-sided) were considered statistically significant. Multivariate logistic regressions were performed to identify the predictors of post-COVID long-term complications. All statistical analyses were performed using Stata/MP 14.0 and GraphPad Prism. The study considered COVID patients showing no symptoms of infection as asymptomatic and those showing at least one of the typical symptoms as symptomatic. Similarly, patients with at least one type of comorbidity were considered comorbid and those with no comorbidity were considered non-comorbid patients.

## Results

### Demographic information and vaccination history of COVID-19 patients in the sample

Of 1,021 COVID-19 inpatients, a total of 111(10.9%) were found to be asymptomatic, while the number of symptomatic cases were 910 (89.1%) which was predominant within the sample (Table-1).

**Table1.**
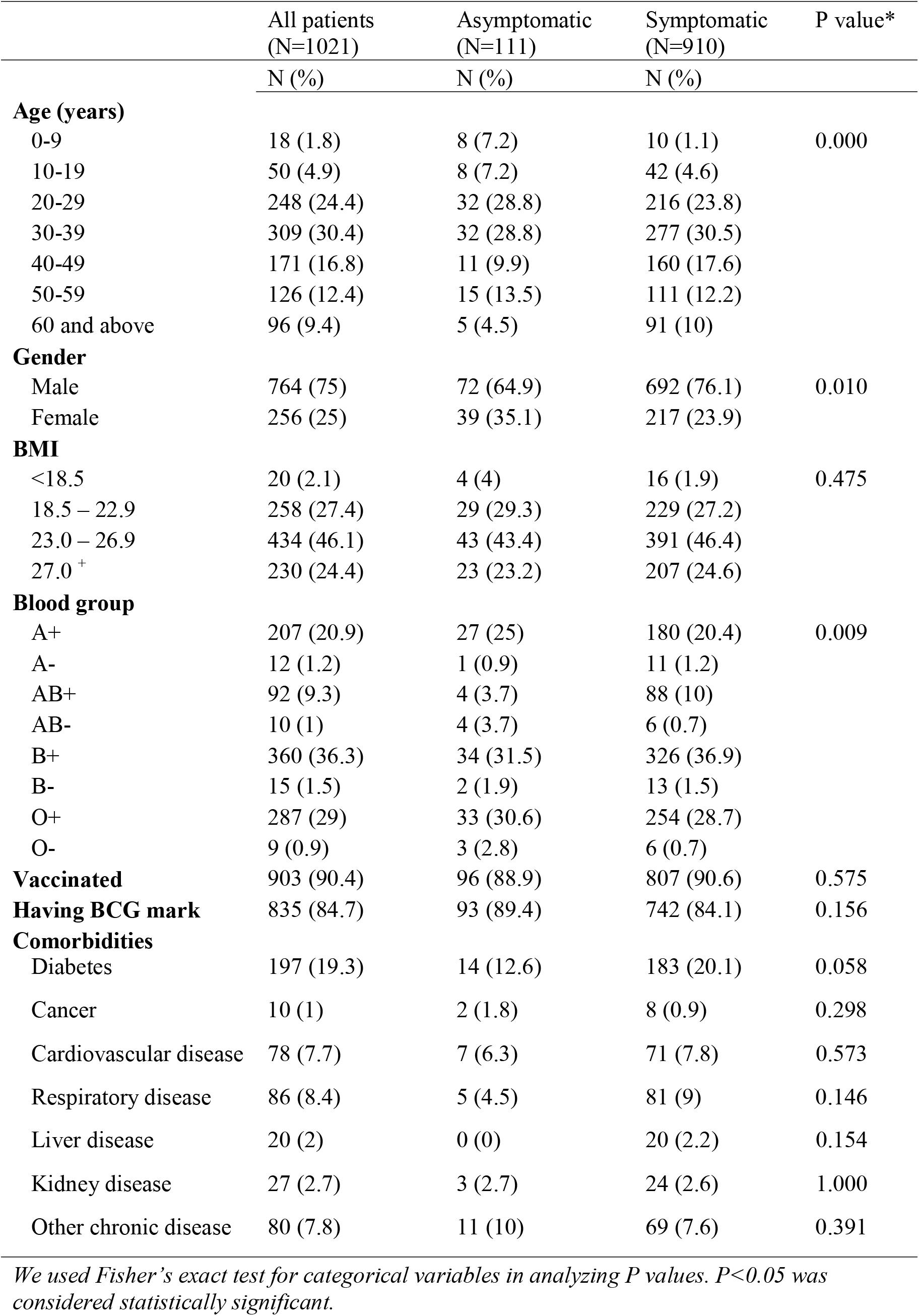
Demographics and baseline characteristics of patients diagnosed with COVID-19

As shown in Table-1, a subject’s age was found to have association with him/her being asymptomatic or symptomatic (p=0.000). Higher proportions of asymptomatic cases were found under the age groups of 0-9 (7.2%), 10-19 (7.2%), and 20-29 (28.8%). Common to both symptomatic and asymptomatic groups, the proportion of males was greater than that of females, the figures being 64.9% and 76.1% respectively (p = 0.01). Individuals in the symptomatic group showed more likelihood towards being overweight (46.4%) and obese (24.6%). A significant bivariate relationship was found (with a p value of 0.009) between blood group and a patient’s being symptomatic or asymptomatic. More than 90% of all patients received all required basic vaccines, and almost 85% had a clear BCG scar on their arms. Diabetes was the most prevalent comorbid condition (19.3%), followed by respiratory (8.4%) and cardiovascular (7.7%) diseases within the total sample. An over the time trend in asymptomatic and symptomatic COVID-19 cases was also examined. The bar graph in Figure-1 clearly illustrates an upward trend among asymptomatic COVID-19 cases with time while the trend for symptomatic cases remained more or less similar.

### Clinical manifestations of COVID-19 patients

Table-2 presents counts and proportions of usual signs and symptoms as observed in individuals diagnosed with COVID-19. Of the total sample, 474 (51%) patients experienced a body temperature exceeding 1010 F, while for 368 (39.5%) patients the body temperature ranged from 990 F to 1010 F. The oxygen saturation rate went below 90% for 19.3% of the patients, and another 8% patient experienced a low saturation level of 90% to 92.9%. The systolic and diastolic blood pressure measures were found to be greater than 159 mmHg and 99 mmHg, respectively for 4.7% of the total cases. The most prevalent symptoms among the cases in order of prevalence rate were fever (72.4%), cough (55.9%), taste loss (40.7%), body ache (40%), smell loss (36.9%), breathing difficulty (25.9%), and sore throat (23.7%).

### Biochemical parameters of COVID-19 patients in the sample

For a substantial number of cases within the sample, biochemical assay results were found to be deviant from the normal range for each of the markers measured in the analyte (Figure-2A). Neutrophils and D-dimer were found to be high for about 50% of the patients in the sample while for approximately 40% of patients, the diagnostic tests showed a high level of Glucose, Ferritin, CRP, and SGPT. Among other parameters, Troponin was high for 30% of the patients, while Creatinine, WBC-TC, and ESR were at elevated levels for around 20% of the patients. A low lymphocyte count was found for 40% of the patients while laboratory test results for 10% of the patients reported low platelet count as well.

### Medication history during the tenure of COVID-19 persistence for cases

Individuals with COVID-19 during the tenure of their SARS-CoV-2 positive status took some day-to-day medications along with antibiotics (Figure-2B). Of the antibiotics, 55.71% of the cases took Azithromycin solely while the number stood at 74% when it was consumed in combination with other therapeutic drugs establishing a higher proportion in terms of intake as compared to the other antibiotics frequently taken. 8.18% of the patients took only Doxycycline and 14.7% took Doxycycline but in combination with other medications, while for Ivermectin the sole consumption and consumption in combination stood at 6.14% and 11.2% respectively. Of the other types of medications, Montelukast was taken by 0.3% and 22.5% of the patients took multiple other types of therapeutic drugs. 6.13% of the patients did not take any medications during COVID-19.

### Preliminary long-herm health issues associated with COVID-19

The most prevalent post-recovery complications that were reported included sleep disturbance (32%), pain and discomfort (31.8%), lack of concentration (24.4%), anxiety and depression (23.1%), memory loss (19.5%), and complications with mobility (17.7%), as shown in Table 3. Considering comorbidity as determinant, chi-square tests for post-recovery complications returned significant deviations between individuals having comorbidity and those without for all reported complications but panic attacks. Comorbid patients were found to be more likely to experience mobility problem (26%), problems performing usual activities (14%), pain and discomfort (40%), anxiety and depression (28.5%), sleep disturbances (41.3%), concentration loss (28.5%), and memory loss (24.6%) than those without any comorbid conditions. We observed significant differences in proportion between the two groups for all complications except panic attacks, giving a clear conception on the comparison between the results of the aforesaid cases with those found for comorbid and non-comorbid cases. Columns in Table-4 contain estimated results from logit models designed for post-recovery complications. As the adjusted odd ratios indicate, the respiratory disease caused each outcome of memory loss and pain and discomfort at 5% level and sleep disturbance at 1%. Finally, it was found that taking drugs for a long time significantly increased the probability of losing memory, concentration, and having sleep disturbance at 1% level and anxiety and depression at 5% level.

**Table 2.** Signs and symptoms of COVID patients

| All patients (N=1021) | N (%) |
| --- | --- |
| **Sign** |  |
| Body temperature |  |
| <99<sup>0</sup> F | 89 (9.6) |
| 99<sup>0</sup> F – 101<sup>0</sup> F | 368 (39.5) |
| >101<sup>0</sup> F | 474 (51) |
| Oxygen saturation rate |  |
| <90% | 80 (19.3) |
| 90%-92.9% | 33 (8) |
| 93%-95.9% | 130 (31.3) |
| >96% | 172 (41.5) |
| Blood pressure |  |
| <120; <80 mmHg | 89 (9.5) |
| 120-140; 80-89 mmHg | 796 (85.2) |
| 141-159; 90-99 mmHg | 5 (0.5) |
| >159; >99 mmHg | 44 (4.7) |
| **Symptom** |  |
| Fever | 738 (72.4) |
| Chills | 127 (12.5) |
| Body ache | 407 (40) |
| Cough | 570 (55.9) |
| Sore throat | 242 (23.7) |
| Breathing difficulty | 264 (25.9) |
| Loss of taste | 415 (40.7) |
| Loss of smell | 376 (36.9) |
| Chest pain | 178 (17.5) |
| Running nose | 190 (18.6) |
| Diarrhea | 202 (19.8) |
| Tiredness | 127 (12.5) |
| Vomiting | 116 (11.4) |
| Blurred vision | 40(4) |

**Table 3.** post-COVID complications of patients by comorbidity status

|  | All patients,<br>(N=1,021) | With<br>comorbidity,<br>(N=358) | Without<br>comorbidity,<br>(N=662) | P value |
| --- | --- | --- | --- | --- |
| Mobility problem | 179 (17.7) | 93 (26) | 86 (13) | 0.000 |
| Problems performing usual activities | 104 (10.7) | 50 (14) | 54 (8.2) | 0.003 |
| Pain and discomfort | 319 (31.8) | 143 (40) | 176 (26.6) | 0.000 |
| Anxiety and depression | 230 (23.1) | 102 (28.5) | 128 (19.3) | 0.000 |
| Sleep disturbances | 312 (32) | 148 (41.3) | 164 (24.8) | 0.000 |
| Panic attack | 121 (12.4) | 43 (12) | 78 (11.8) | 0.712 |
| Lack of concentration | 243 (24.4) | 102 (28.5) | 141 (21.3) | 0.005 |
| Memory loss | 195 (19.5) | 88 (24.6) | 107 (16.2) | 0.001 |

**Table 4.** Adjusted odds ratios for predictors of post-COVID complications

|  | 1 | 2 | 3 | 4 | 5 | 6 | 7 |
| --- | --- | --- | --- | --- | --- | --- | --- |
|  | Memory loss | Concentration inability | Anxiety & depression | Mobility problem | Activity problem | Sleep disturbance | Pain & discomfort |
| Diabetes | — | — | — | — | — | — | — |
| Cancer | — | — | 0.10*<br>(0.138) | — | — | — | — |
| Cardiovascular disease | — | — | — | — | — | — | — |
| Respiratory disease | 1.70**<br>(0.477) | 1.62*<br>(0.433) | — | — | — | 2.41***<br>(0.652) | 1.70**<br>(0.445) |
| Kidney disease | — | — | — | — | — | — | — |
| Liver disease | — | — | 9.76***<br>(6.643) | 6.06***<br>(3.499) | 5.07***<br>(2.957) | 2.88*<br>(1.701) | — |
| Other chronic disease | — | — | 1.95**<br>(0.546) | — | — | — | — |
| Prolonged medication use | 1.85***<br>(0.436) | 1.84***<br>(0.403) | 1.65**<br>(0.375) | — | — | 2.16***<br>(0.470) | — |
| No. of observations | 902 | 898 | 894 | 905 | 863 | 872 | 901 |

All models are controlled for age, gender, BMI, vaccination. \*\*\* P<0.01, \*\* P<0.05, \* P<0.10, Parentheses include standard errors. Coefficients are reported if they are statistically significant

### Contact History of the COVID-19 Patients in this Sample

This study also assessed the contact history of patients included in the study sample, focusing on multiple variables representing direct and indirect contacts (Supplementary file, Table-S1). As Table S1 shows 50% of the diagnosed cases came in close contact with other COVID-19 positive individuals, while 40.6% patients of the total sample had family members diagnosed with COVID-19. A total of 608 (62%) patients reported to have an indirect contact, and 466 (48.5%) patients made frequent visits outside their home prior to being infected.

## Discussion

In the current study, we investigated 1021 COVID-19 inpatients diagnosed through RT-PCR in six different health facilities in Bangladesh. We believe that the current study will be able to shed light on the epidemiological aspects of COVID-19 during the crisis period. While the study revealed facts aligning with those established globally some observations require further follow up and extensive studies in Bangladesh.

Our study has revealed that among the age groups, the 20-39 cohorts showed the highest prone to infection rate (54.7%) and in terms of gender, the prevalence of SARS-CoV-2 infection in males (74.9%) was 3 times more than that in females (25.1%). Several studies from across the globe showed higher median age,^9-11^ mean age,^12^ or age group^9^ for SARS-CoV-2 infection. We explained the observations in light of demographic and socio-economic situations in Bangladesh. 21-40 years is the most preponderant age group in Bangladesh (https://www.cia.gov/library/publications/the-world-factbook/fields/341.html), young and middle-aged people mostly fall under the working class and are needed to go are mostly obligated to leave home for work, and hence having higher odds of getting infected.^13^ Females, especially in rural Bangladesh are mostly confined to household work, which might be a cause for less infection among these groups.

Among the study subjects, 38.33% had comorbidities. Diabetes was found to be the most prominent comorbidity (19.40%) within the study sample and this fact was reflected in similar studies conducted in China and UK.^11, 14^ Other noteworthy comorbidities were respiratory (8.40%) and cardiovascular diseases (7.70%) and the results obtained from our study aligned to the trends in other countries in terms of high prevalence.^9, 10, 14^

As for the symptoms associated with COVID-19, fever (72.4%) and cough (55.9%) were the most prominent and shared among cases (Table-2). Other clinical manifestations with significant association (P<0.01) were breathing difficulty (25.90%) and chills (12.50%). Diarrhea and GI symptoms were also widely observed in 19.80% cases of COVID-19 in the study sample akin to results obtained from previous studies conducted on the MERS/SARS-CoV pandemics.^15^

Of the 1,021 COVID-19 positive cases in our sample, 10.9% were asymptomatic which is in accordance with the findings in South Korea and China.^9, 16^ Interesting enough, our observation identified an upward trend for the increase in asymptomatic COVID-19 cases following April 2020 (Figure 1). Although no systematic assessment has been done on this aspect, our observation complies with the records and pronouncements of the health professionals and hospital registries in Bangladesh. For getting a better insight on the upward trend of asymptomatic COVID-19 cases further studies and scrutinization are required including a larger sample and dataset.

**Figure 1.**
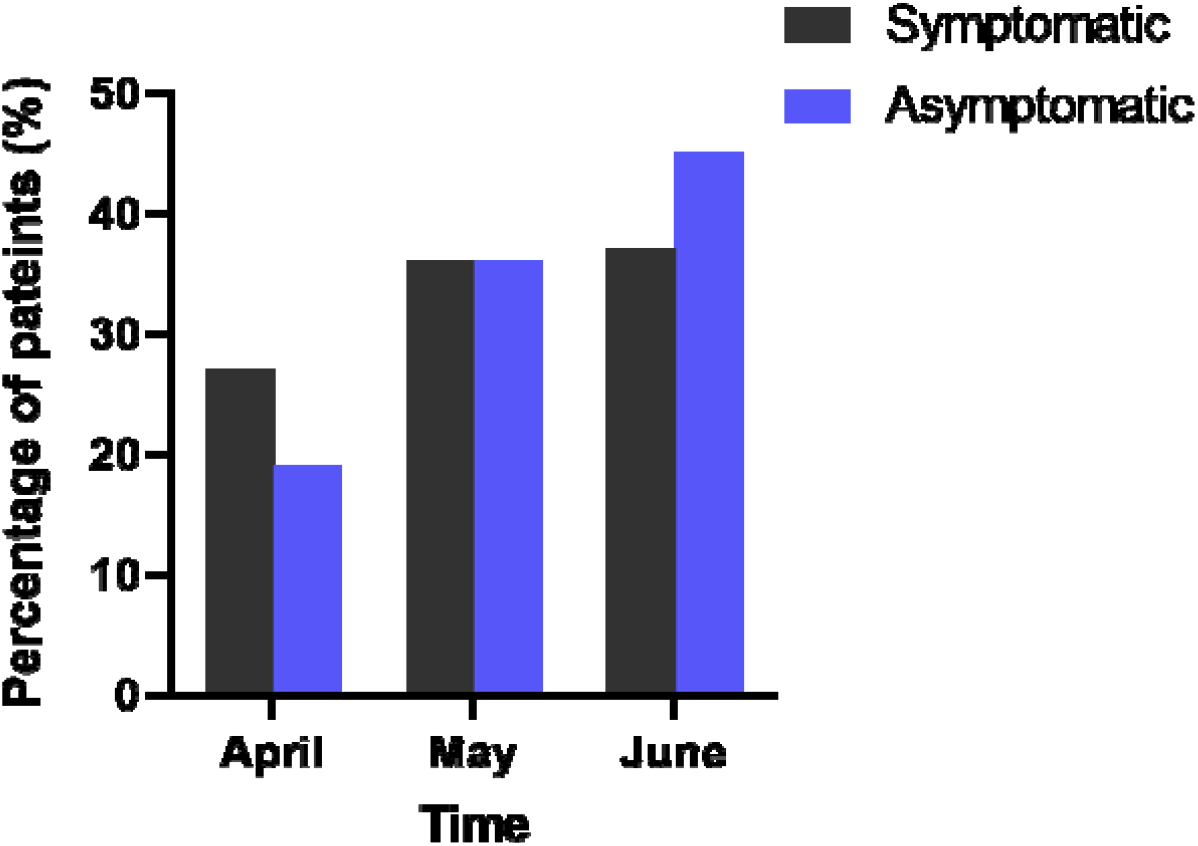
Frequency of symptomatic and asymptomatic COVID-19 patients at various time points.

The current study observed different parameters obtained from biochemical assays conducted within the sample and categorized the results as low, normal and high compared to a reference value (Figure 2A). Almost half of the cases had a high level of D-dimer (46.12%) establishing COVID-19 related coagulopathy which is likely a manifestation of profound inflammatory response.^17^ For the other parameters, rise in ferritin, CRP, ESR and glucose levels were found in 37.92%, 36.08%, 27.37% and 36.48% of the COVID-19 cases respectively. These findings were found to comply with results obtained from similar studies in China conducted on a sample size of 99 and 138 patients respectively.^18, 19^

**Figure 2.**
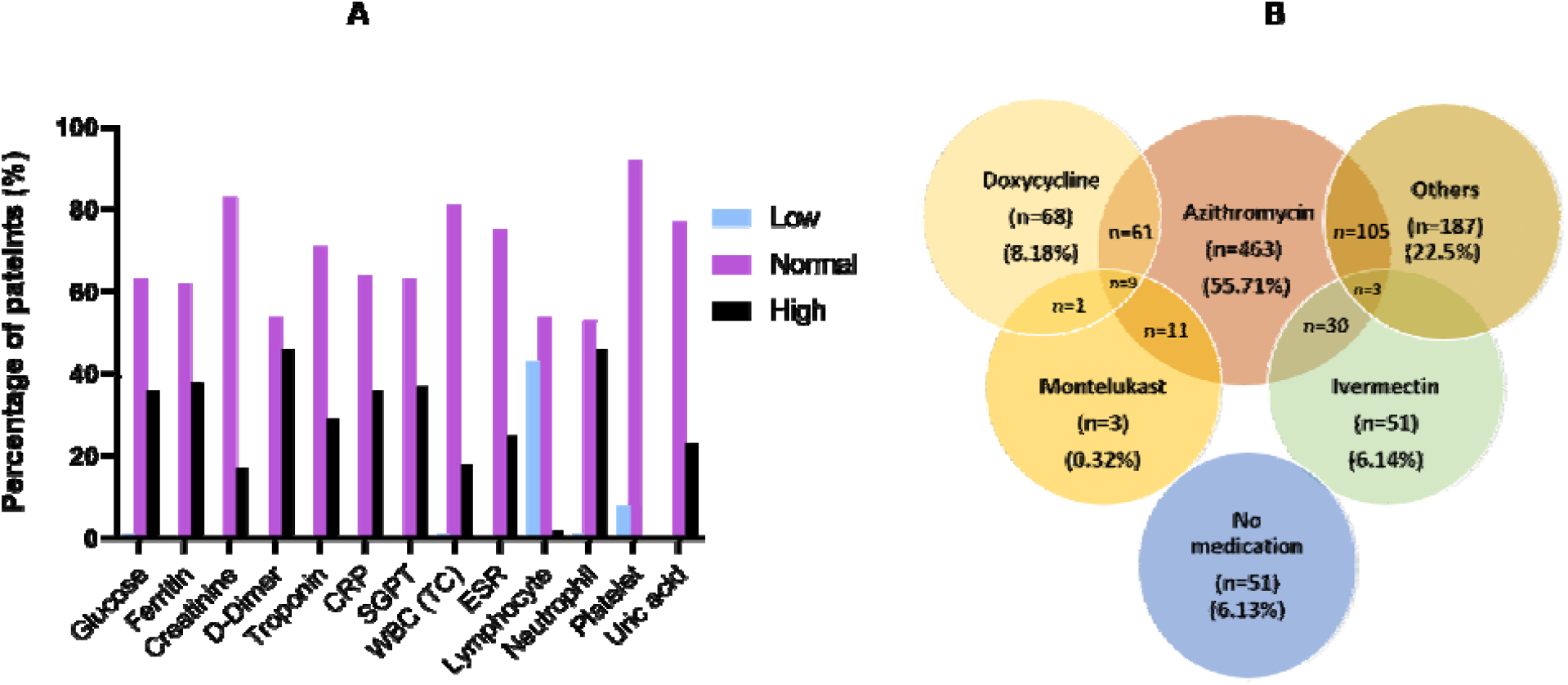
Biochemical parameters and medication history of COVID-19 patients. A) Various biochemical measurements during onset of COVID-19. Normal reference values are: D-dimer < 0.5 µg/mL, Ferritin-Male 18-370 ng/mL, Female 9- 120ng/mL; Serum Creatinine-Male 0.4-1.4 mg/dL, Female - 0.3mg −1.1mg/dL; CRF: < 5mg/L; Troponin: < 0.30 ng/mL; Uric acid: Male- 3.4 – 7 mg/dL, Female 2.4 – 6 mg/dL; ESR: Male < 22 mm/hr, Female < 29mm/hr; SGPT: Male: 15-65 U/L, Female 15 −60 U/L; CBC : WBC: 4-11 x 109/L, Neutrophil: 40-75%; lymphocyte 20-45%, ESR: male 0-10 mm, Female 0-15 mm. B) Medication history of the patients during the tenure of COVID-19 persistence for cases.

Our study revealed that 28.75% cases of patients had raised troponin level and it is assumed that this is owing to myocarditis, microangiopathy, myocardial infarction, cytokine storm etc.^17, 20-23^ Elevated liver enzyme, SGPT was also found in 36.83% cases in this study which is also in accordance with in many other papers confirmed by review.^24^ Although, possible mechanism is yet to be confirmed, majority of these cases reported use of multiple drugs like antibiotics, antivirals, hydroxychloroquine and steroids, which could have contributed to the liver cell damage. Additionally, ischemia and immune mediated injury might be responsible for elevated liver enzyme in Covid-19 positive cases.^24^ These observations suggest that patients diagnosed with COVID-19 should also undergo follow up diagnosis procedures to check for chronic damages of the heart and liver.

Total cell count reports for COVID-19 positive individuals indicated that neutrophil count was elevated in 46.44% of the cases. On the contrary, reduced lymphocyte and platelet counts have been observed among 50% and 10% COVID-19 positive patients. This is in accordance with several other case series around the globe.^17, 25-28^ As our sample included individuals recovered from COVID-19, it is clear that these fluctuating parameters for the blood cell counts did not become lethal to the patient. However, whether they were related to any severe symptom previous to recovery requires further investigation. Our study did not investigate the level of cytokines or other inflammatory mediators in the patients. Further studies in this regard will be helpful in understanding the severity and fatality among COVID-19 patients with respect to immune response mediators.

Human-to-human transmission of SARS-CoV-2 has been established as the major mode of spread and several reports provided evidence of transmission of COVID-19 by direct and indirect contact in hospital, family and community settings where interventions were not applied and basic health norms and distancing were not maintained.^29-32^ In our study, majority of the cases (62%) reported indirect contact with confirmed cases, whereas, 48.5% admitted to go out of their homes frequently prior to being infected and diagnosed (Table-S1, supplementary). It further shows that 50% of the respondents had close contact with confirmed cases and 40.6% had COVID-19 positive family members. So, it can be easily concluded from this data that community transmission is very common in Bangladesh and hence social distancing and other preventive measures should be fully implemented to prevent further deterioration.

Our study has revealed that antibiotics were heavily suggested as a part of the treatment protocol for the COVID-19 patients (Figure 2B). It has been found that Azithromycin was the most prescribed antibiotic (74%); among them 55.71% were prescribed to take it as a sole drug. Doxycycline and Ivermectin were taken by 14.7% and 11.2% cases respectively; among them 8.18% and 6.14% patients got them exclusively. Supportive drugs like montelukast were also taken by a very small number of patients (3%). In Bangladesh, the guideline for COVID-19 management allows compassionate use of various antiviral and anti-bacterial drugs along with antimalarial drugs (https://dghs.gov.bd/index.php/en/). However, we recommend that the excessive use of Azithromycin and other antibiotics may aggravate the crisis related to the rise of antibiotic resistance in Bangladesh.

Whether BCG vaccination may play a significant role in preventing the SARS-CoV-2 infection has remained an area of debate and requires further analysis and assessment. According to the latest WHO brief,^33^ BCG does not give protection against SARS-CoV-2 infection. In our study, we have found that 835 (82%) cases have a BCG vaccination mark (Table-1). Therefore, supporting the WHO brief, we infer that BCG vaccination may not play any role in preventing SARS-CoV-2 infection despite some early speculations about it. However, as the subjects are all alive despite SARS-CoV-2 infection, it will be insightful to further investigate whether BCG vaccination may help reduce the severity and hence mortality by modulating the innate immune system.^34^

Our study has also shed light on the relationship between the ABO blood groups and susceptibility to SARS-CoV-2 infection. A number of previous reports suggested individuals having an A (+ve) blood group have the highest risk of being infected while O (+ve) individuals have a comparatively lower risk rate.^35-37^. Our study has showed that the majority of the cases belong to B+ve blood group (35.4%), preceded by O (+ve) 28.2% and A (+ve) 20.3% (Table-1). This ratio is comparable with the population distribution of ABO blood groups (B+ve ∼34%, O+ve ∼30% and A+ve ∼26%) in Bangladesh.^38^ So, we assume that the variation in the geographical prevalence of different blood groups might be the reason behind the findings of certain ABO blood groups being more common in certain countries without having any real contribution to COVID-19 susceptibility.

An important part of our study was to assess and scrutinize the sample for post-COVID complications-both physical and mental (Table-3 and Table-S2, supplementary file). Our study found that people with comorbidities have reported post-COVID complications such as mobility problems, pain and discomfort, anxiety and depression and indication of memory loss with a greater significance (Table-4). We also found that the vulnerability of recovered individuals towards post COVID complications was due to being comorbid and being exposed to drugs for a prolonged period (Table-4). Although retrieved from a basic questionnaire, the results obtained from this study are statistically significant. Our findings are consistent with data obtained from similar studies in other countries.^39, 40^

Since this study was an onset in regards to epidemiological analysis of COVID-19 patients in Bangladesh, it was prone to some limitations. Firstly, the study sample only included inpatients from 6 selected hospitals providing aid to COVID-19 cases in Bangladesh which narrows down the sample size and diversification in results. Secondly, since the data were collected over the telephone there may be possibilities of information bias, interviewer bias as well as recall bias. Finally, commenting on an individual’s health status and complications post COVID-19 depending on a four-week recovery period is prompt. Nevertheless, this study aimed to observe the early post-COVID-19 health complications.

To date this is the most comprehensive study conducted in Bangladesh that assessed COVID-19 patients for both pre-recovery and post-recovery complications. Our data have reconfirmed some significant global observations in regards to the epidemiological and clinical aspects of the disease, but above all, the study has helped in addressing concerns that raised eyebrows including the role of ABO blood group and BCG vaccination in COVID-19 susceptibility and progression. We anticipate that the outcomes of this study will work as a baseline for future studies in the same context.

## Conclusion

To sum up, this study identified the early health complications post recovery in COVID-19 patients which yet has aspects that require further assessment and examining for more in-depth insights. This study found that individuals with comorbidities had a hard time coping with the clinical implications imposed while they were with COVID-19. The fact that there is a significant advancement of the number of asymptomatic COVID-19 cases calls for further investigations. The results obtained from this study could contribute to devising effective strategies for the provision of comprehensive health care to COVID-19 patients with comorbidity.

## Data Availability

The authors agree.

## Acknowledgements

Authors would like to thank the volunteers and research assistants of Disease Biology and Molecular Epidemiology Research Group, Chittagong for their help and contribution to complete this study.

